# Multimodal Machine Learning for Glaucoma Detection in a Sub-Saharan African Clinical Population

**DOI:** 10.64898/2026.03.13.26347955

**Authors:** Emmanuel Adator, Andrew Owusu-Ansah, Mercy Oforiwaa Berchie, Julius Markwei, Joseph Sa-Ambo, Kwame Anag-bey, Abraham Yiadom Boakye, Samuel Kyei, Enyam Morny, Emmanuel Addai

## Abstract

**Purpose:** To evaluate the performance of machine learning models for automated glaucoma detection using multimodal clinical, structural, and functional data from a West African clinical cohort.

**Methods:** In this retrospective observational study, we analyzed clinical records from two major eye care centers in Ghana. A total of 605 eyes from 417 patients who underwent comprehensive glaucoma evaluation were included. Extracted features included demographic data, intraocular pressure, optical coherence tomography (OCT) structural parameters, and Humphrey visual field indices. We assessed the diagnostic performance of individual parameters using receiver operating characteristic (ROC) analysis. Supervised machine learning classifiers, including support vector machine (SVM), random forest (RF), gradient boosting machine (GBM), and a multi-layer perceptron (MLP), were trained using a forward feature selection approach and evaluated using five-fold cross-validation. We assessed model performance by computing performance metrics like sensitivity, specificity, and area under the ROC curve (AUC).

**Results:** Among the 605 eyes analyzed, 361 (59.7%) were glaucomatous, and 244 (40.3%) were healthy. Individual structural and functional parameters demonstrated moderate discriminative ability, while age showed no significant diagnostic value (AUC = 0.49, p = 0.841). Among machine learning models, the MLP achieved the highest diagnostic performance (AUC = 0.90 [95% CI: 0.86–0.92], sensitivity = 0.88, specificity = 0.86), outperforming SVM (AUC = 0.82), RF (AUC = 0.78), and GBM (AUC = 0.77).

Multimodal integration of clinical, structural, and functional features improved discrimination compared with individual parameters.

**Conclusions:** Multimodal machine learning models can effectively automate glaucoma detection using routinely collected clinical data. In this West African cohort, an MLP-based approach demonstrated superior diagnostic performance compared with traditional machine learning models and individual clinical metrics. These findings highlight the potential of clinically grounded artificial intelligence tools to support glaucoma diagnosis and triage in resource-constrained eye care settings.

## INTRODUCTION

Glaucoma is the commonest cause of irreversible blindness worldwide and disproportionately affects people of African descent [1], [2], [3], [4], [5]. These populations experience earlier disease onset, more aggressive progression, and higher rates of undiagnosed disease compared to other populations [6], [7], [8]. In West Africa, late presentation and limited access to subspecialty eye care further exacerbate visual morbidity, underscoring the need for scalable, accurate, and context-appropriate diagnostic tools [9], [10]. Despite advances in imaging and functional testing, glaucoma diagnosis continues to rely on expert interpretation of heterogeneous clinical data, which is both time-intensive and subject to inter-clinician variability [11]. This diagnostic burden is particularly pronounced in low-resource settings such as sub-Saharan Africa, where clinicians often manage high patient volumes with limited specialist workforce and few structured systems for triaging high-risk patients [7].

Advances in machine learning (ML) have generated considerable interest in automating glaucoma detection using multimodal ophthalmic data [12], [13], [14]. Evidence from multiple studies indicates that retinal imaging, OCT, VFT indices, and routine clinical parameters provide reliable diagnostic performance, often reporting high accuracy and area-under-the-curve metrics [15], [16]. However, many of these models are developed and validated using datasets derived predominantly from European, East Asian, or mixed populations, with minimal representation of African eyes [17]. This raises critical concerns about the generalizability, fairness, and clinical validity of such models when applied to populations with distinct ocular anatomy, disease phenotypes, and healthcare contexts [17], [18], [19].

Emerging evidence suggests that structural and functional manifestations of glaucoma may differ systematically for populations of African descent, including variations in optic disc size, retinal nerve fiber layer (RNFL) thickness, and structure–function relationships [6], [20]. Consequently, ML models trained on non-African datasets may underperform or behave unpredictably when deployed in African clinical settings, potentially reinforcing existing healthcare disparities rather than mitigating them [12], [18], [19]. Despite these risks, population-specific evaluations of ML-based glaucoma diagnostics in sub-Saharan Africa (SSA) remain absent.

Beyond population bias, many ML glaucoma studies rely on highly curated datasets, ideal imaging conditions, or single-modality inputs that do not reflect routine clinical practice in low-income countries [19], [21]. In real-world SSA clinics, diagnostic data are often heterogeneous and acquired under resource constraints, yet these conditions represent the true environment in which automated tools are expected to function. There is, therefore, a critical need for ML approaches that are not only accurate but also robust and clinically grounded within the realities of SSA eye care systems.

In this context, the present study investigates the automation of glaucoma diagnosis using supervised machine learning techniques applied to a West African clinical cohort. By leveraging routinely collected clinical, structural, and functional diagnostic data and linking model development to clinically established definitions of glaucoma, this work evaluates the diagnostic performance of ML–assisted glaucoma detection in a population that has been historically underrepresented in ophthalmic artificial intelligence research. Beyond comparative model performance, the study also considers the potential implications of such approaches for real-world clinical deployment in West African eye-care settings, where resource constraints and specialist shortages remain significant challenges.

## METHODS

### Research Design

We used a retrospective observational study approach and collected clinical records from two tertiary eye care facilities in Ghana: the Bishop Ackon Memorial Eye Center and the Eye Clinic of the School of Optometry and Vision Science at the University of Cape Coast (UCC). These facilities function as referral centers and deliver comprehensive ophthalmic services to patients drawn from a wide range of communities and healthcare levels.

We evaluated the applicability and diagnostic ability of machine learning (ML) algorithms for automated glaucoma detection using routinely collected clinical and demographic information. By utilizing real-world patient records rather than highly curated research datasets, the intention was to develop models that reflect practical diagnostic conditions, particularly in settings with limited resources.

We obtained approval from the ethical review committee of UCC (Ethical Clearance: UCCIRB/CHAS/2024/06). Before analysis, all patient information was de-identified to maintain confidentiality. We conducted the study in accordance to institutional guidelines governing data protection.

### Study Population

We included records of patients who had undergone comprehensive glaucoma evaluation and were diagnosed by ophthalmologists as either glaucomatous or non-glaucomatous. These labels served as the reference standard for model training and validation. Records were included if they contained complete data from the same diagnostic period, including Optical Coherence Tomography (OCT; RTVue) structural parameters, Visual Field Test (VFT) indices, intraocular pressure (IOP), corrected visual acuity, and age.

We excluded records with missing modalities, poor-quality OCT or unreliable VFT, incomplete diagnostic labels, or coexisting ocular diseases that could confound glaucoma assessment. This ensured all included data were clinically valid and consistently labeled.

### Data Acquisition and Feature Representation

We extracted structural, functional, and clinical data retrospectively from patient records. OCT measurements provided structural parameters, visual fields provided functional indices, and examination records supplied clinical and demographic data.

We included only routinely collected, quantitative parameters to maintain consistency and interpretability. Feature selection was guided by clinical relevance. Each record was assigned a unique anonymized identifier before analysis. A summary of all variables extracted from patient medical records is listed in Table 1 and categorized according to subthemes.

**Table 1.** Summary Features Extracted from Patient Medical Records.

| Domain | Variable | Description |
| --- | --- | --- |
| Demographic | Age | Patient age at time of examination (years) |
| Clinical Examination | Best-corrected visual acuity (BCVA) | Visual acuity after refractive correction |
|  | Intraocular pressure (IOP) | Measured intraocular pressure |
| OCT – Retinal Nerve Fiber Layer (RNFL) | Mean RNFL thickness | Average RNFL thickness across the peripapillary region |
|  | Superior RNFL | RNFL thickness in the superior quadrant |
|  | Inferior RNFL | RNFL thickness in the inferior quadrant |
|  | Nasal RNFL | RNFL thickness in the nasal quadrant |
|  | Temporal RNFL | RNFL thickness in the temporal quadrant |
|  | RNFL sectoral values | Sector-specific RNFL measurements |
| OCT – Optic Nerve Head (ONH) | Disc area | Total optic disc area |
|  | Neuroretinal rim area | Area of remaining neural rim tissue |
|  | Vertical cup-to-disc ratio | Ratio of vertical cup diameter to disc diameter |
|  | Horizontal cup-to-disc ratio | Ratio of horizontal cup diameter to disc diameter |
|  | Cup volume | Volume of the optic cup |
|  | Rim volume | Volume of the neuroretinal rim |
|  | Cup area | Surface area of the optic cup |
|  | Nerve head volume | Total optic nerve head volume |
| OCT – Ganglion Cell Complex (GCC) | Mean GCC thickness | Average thickness of the ganglion cell complex |
|  | Superior GCC | GCC thickness in the superior region |
|  | Inferior GCC | GCC thickness in the inferior region |
|  | Global loss volume (GLV) | Diffuse GCC loss index |
|  | Focal loss volume (FLV) | Localized GCC loss index |
| Visual Field Metrics | Mean deviation (MD) | Overall visual field loss (dB) |
|  | Pattern standard deviation (PSD) | Localized field irregularity (dB) |
| Target Variable | Glaucoma classification | Diagnostic label (0 = non-glaucoma, 1 = glaucoma) |

### Data Preprocessing

All extracted variables were first examined for missing data, outliers, and values that appeared clinically implausible before proceeding with model development. We excluded eyes that had more than 20% missing data across the selected features. We also evaluated overall data quality using established clinical and imaging reliability indicators, including VFT reliability indices and OCT signal strength. Records that did not meet the predefined quality criteria were removed from the dataset.

For the remaining data, we addressed missing values in the training set through mean imputation. We standardized continuous variables using z-score normalization. Normalization parameters were calculated for the training data and subsequently applied to the test sets to ensure consistency during model evaluation.

### Diagnostic Performance of Individual Clinical Parameters

To contextualize the performance of our models, we evaluated the diagnostic ability of individual demographic, clinical, structural (OCT), and functional (VFT) parameters in identifying healthy from glaucomatous eyes in comparison to the ability of our models in identifying glaucoma. We evaluated the diagnostic performance of each parameter by computing the area under the receiver operating characteristic (AUROC) curve and the confidence interval at 95%.

We determined optimal operating points for sensitivity and specificity using the Youden Index (J = sensitivity + specificity − 1), which is the threshold that maximizes the combined diagnostic yield of each parameter. Statistical significance of glaucoma detection was assessed using ROC-based hypothesis testing. Parameters with non-significant AUROC values were interpreted as having limited standalone diagnostic utility. We used this analysis as the benchmark for comparing the performance of individual clinical metrics against our models trained on combined demographic, clinical, structural, and functional features.

### Machine Learning Model Development

We implemented supervised ML models to classify eyes as glaucomatous or non-glaucomatous based on clinical labels. We compared three traditional classifiers: support vector machine (SVM), random forest (RF), and gradient boosting machine (GBM). The data was additionally trained on a multi-layer perceptron (MLP).

The MLP consisted of two hidden layers (64 and 34 neurons) (Figure 1). We designed the network to capture non-linear relationships while minimizing overfitting. We optimized model parameters using the adaptive moment estimation (Adam) optimization algorithm, which updates network weights through adaptive estimates of first and second moments of the gradients to facilitate efficient and stable training [22]. The network was designed to predict the probability of an eye being glaucomatous, with class labels encoded as 0 for non-glaucoma and 1 for glaucoma. The final output node used a sigmoid activation function to generate probability estimates bounded between 0 and 1. A decision threshold of 0.5 was applied, such that predicted probabilities equal to or greater than 0.5 were classified as glaucoma, while values below this threshold were classified as non-glaucoma.

**Figure 1.**
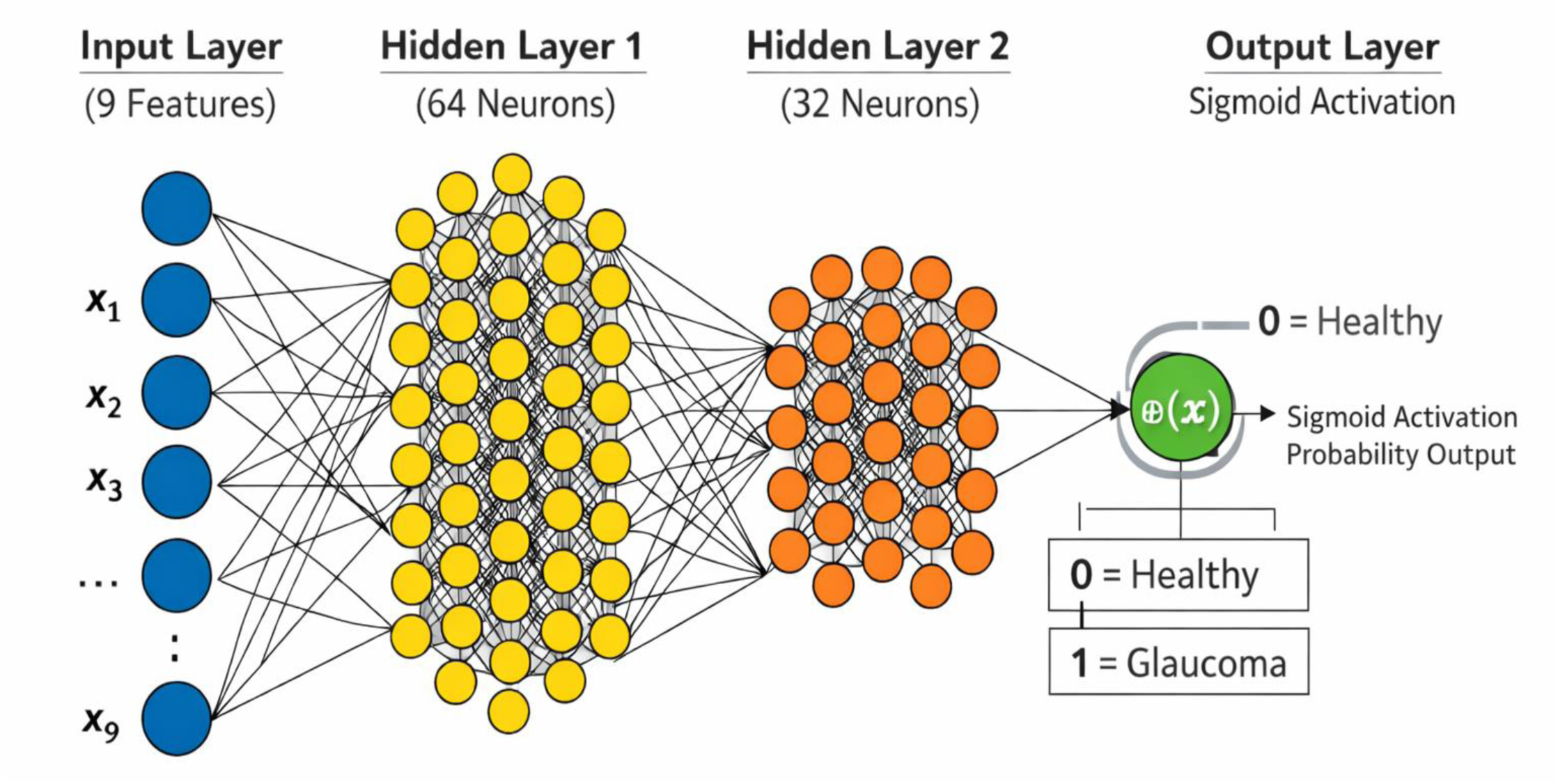
The MLP model consists of: (a) an input layer with nine features (age, intraocular pressure, optic disc area, MD, GLV, FLV, mean GCC, superior GCC, and inferior GCC thickness), (b) a hidden layer containing 64 and 32 sub-layered neurons and (c) an output layer which produces a binary classification (0 = healthy, 1 = glaucoma) using a sigmoid activation function

Model development prioritized predictive performance and clinical relevance. We used a forward feature selection technique to identify the smallest subset of clinically meaningful features that optimized diagnostic performance (Table 2). For all four models, that is, the SVM, RF, GBM, and MLP, we implemented a Five-fold cross-validation technique to guide data partitioning, training, and evaluation. At the beginning of model development, the complete dataset was randomly split into five roughly equal folds. For each iteration, four folds (representing 80% of the data) were used for model training, while the remaining fold (20%) was reserved solely for validation. This procedure was repeated five times so that each fold served once as the validation set.

**Table 2.** Features Selected for Model Training Using Forward Feature Selection.

| Data Domain | Variable |
| --- | --- |
| Demographic | Age, years |
| Clinical Examination | Intraocular Pressure (IOP), mmHg |
| OCT Structural – Optic Nerve Head | Disc Area, mm<sup>2</sup> |
| Visual Field Assessment | Mean Deviation (MD), dB |
| OCT Structural – Ganglion Cell Complex | Global Loss Volume (GLV), % |
|  | Focal Loss Volume (FLV), % |
| | Mean GCC Thickness, μm |
| | Superior GCC Thickness, μm |
| | Inferior GCC Thickness, μm |

Within each cross-validation iteration, all preprocessing steps, including feature selection, imputation, scaling, and model fitting, were performed using only the training folds. The trained model was subsequently applied to the corresponding held-out validation fold, which remained completely unseen during model training and preprocessing, to generate predictions for performance evaluation. At no point were validation data used to inform feature selection, preprocessing parameters, or model training, thereby preventing information leakage. This same cross-validation structure was applied uniformly across all classifiers to ensure direct and fair comparison of performance.

For each model, performance metrics were computed separately for each validation fold and subsequently averaged across all six folds to obtain final estimates of sensitivity, specificity, and area under the receiver operating characteristic curve.

### Model Evaluation

Model diagnostic ability was assessed using sensitivity, specificity, and AUROC. Sensitivity quantified the capacity of models to correctly identify glaucomatous eyes, whereas specificity quantified the capacity of models to correctly detect healthy eyes. The AUROC provided a summary measure of the overall discriminative performance of the models. Evaluation metrics were calculated within the cross-validation framework and reported as mean values across the validation folds.

## RESULTS

605 eyes of 417 patients who had attended the Bishop Ackon Memorial Eye Center and the University of Cape Coast School of Optometry Eye Clinic were used in the final modelling and analysis. All included records were for subjects who had undergone a comprehensive glaucoma assessment and had been clinically diagnosed by licensed ophthalmologists or optometrists as either being healthy or having glaucoma.

Of the 605 eyes analyzed, 361 (59.7%) were diagnosed with glaucoma, while 244 (40.3%) were classified as healthy. The study population comprised 367 (60.7%) male and 238 (39.3%) female eyes.

Baseline characteristics and summary statistics of selected diagnostically relevant structural, functional, and clinical parameters for the glaucoma and non-glaucoma groups are presented in Table 3. Clinically meaningful differences were observed between groups across key parameters commonly used in glaucoma evaluation.

**Table 3.** Baseline Characteristics of Patients with Healthy and Glaucoma Groups.

| Variable | Healthy Group | Glaucoma Group | P-Value |
| --- | --- | --- | --- |
| Total eyes (%) | 361 (59.7%) | 244 (40.3%) | - |
| Gender (Male/Female) | 233 (141/92) | 372 (226/146) | = 0.95 |
| Age (years) | 39.12 (± 12.83) | 51.09 (± 14.51) | < 0.001 |
| Mean deviation (dB) | -1.74 (2.91) | -10.71 (10.02) | < 0.001 |
| Average RNFL (μm) | 112.40 (± 8.26) | 88.82 (± 18.30) | < 0.001 |
| Average GCC (μm) | 98.77 (±6.02) | 81.68 (±12.70) | < 0.001 |

Demographic, clinical, structural, and functional characteristics of the study population are presented according to disease state. Numerical data are reported as mean ( ± standard deviation), while categorical variables are reported as number (percentage).

The diagnostic performance of individual clinical, structural, and functional parameters is summarized in Table 4. Most single parameters demonstrated moderate discriminative ability, with considerable variability in sensitivity–specificity trade-offs. Age alone showed no significant diagnostic value (p =0.841).

**Table 4:**
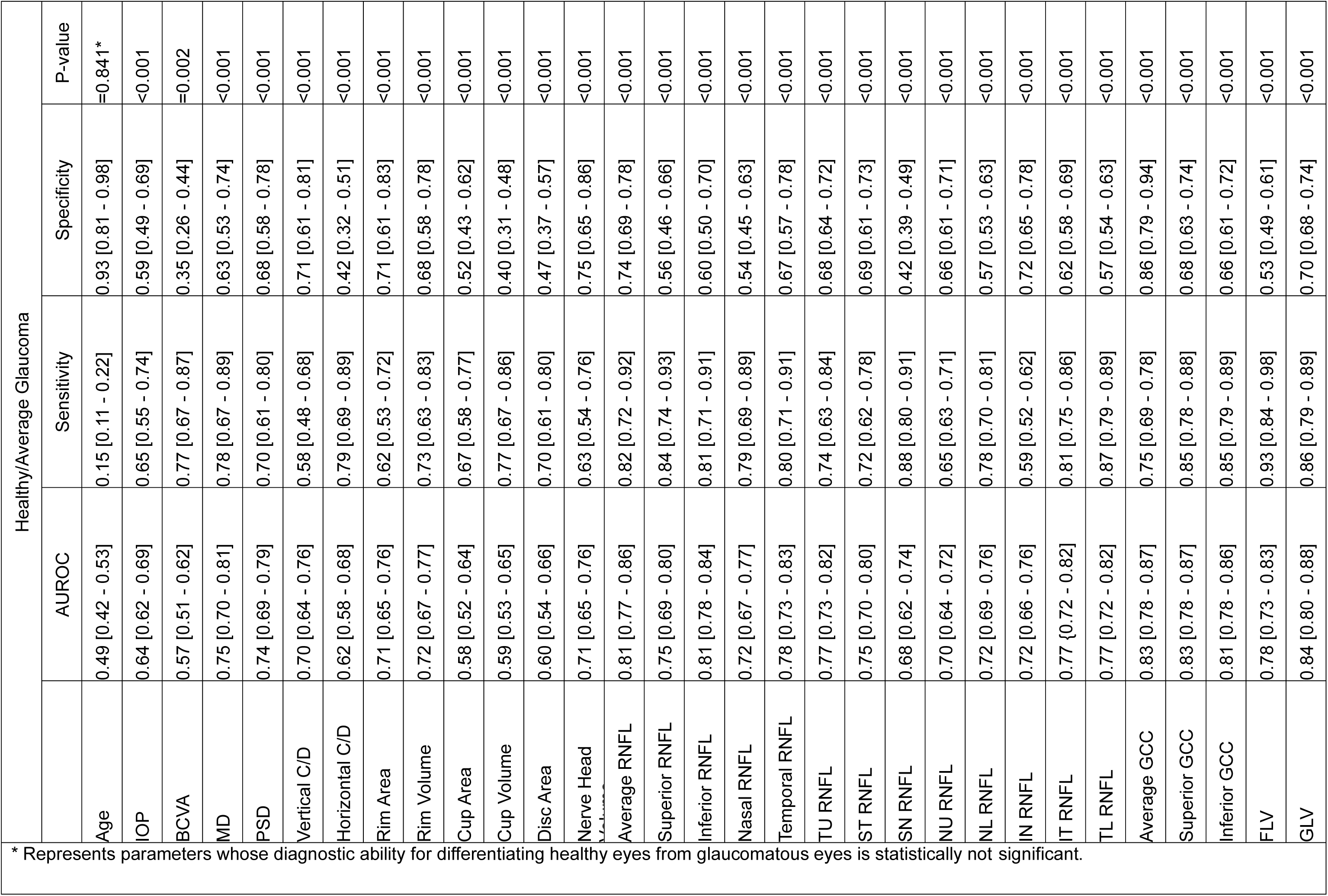
Area under the ROC curve (AUROC) and Optimal Sensitivity–Specificity Balance of Parameters in Differentiating Healthy Eyes from Glaucomatous Eyes.

All models demonstrated good discriminatory ability for glaucoma, with variations in performance observed across classifier types (Figure 2). Traditional ML classifiers, including SVM, RF, and GBM, showed comparable performance, while the MLP achieved competitive discrimination when trained on the selected feature subset. Overall model performance metrics across all classifiers are summarized in Table 5.

**Figure 2.**
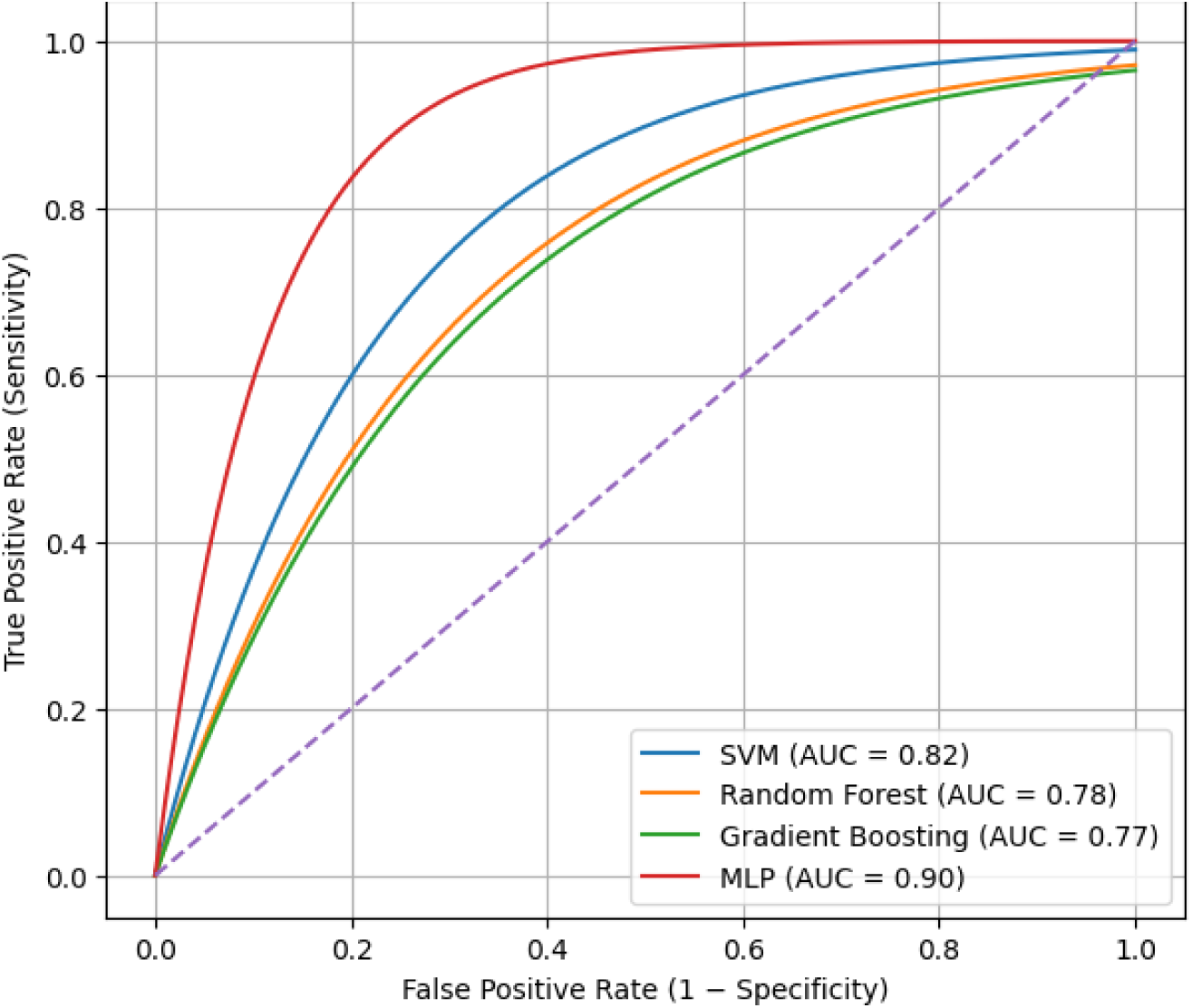
AUROC curves showing the relationship between the true positive rate and false positive rate for each model alongside their computed diagnostic ability.

**Table 5.** Performance of Machine Learning Models for Glaucoma Detection.

| Model | Sensitivity (%) | Specificity (%) | AUC | P-value |
| --- | --- | --- | --- | --- |
| Support Vector Machine (SVM) | 82 [78–86] | 79 [74–83] | 0.82 [0.79–0.84] | <0.001 |
| Random Forest (RF) | 80 [73–83] | 71 [68–74] | 0.78 [0.75–0.81] | <0.001 |
| Gradient Boosting Machine (GBM) | 76 [72–79] | 69 [64–72] | 0.77 [0.74–0.79] | <0.001 |
| Multi-Layer Perceptron (MLP) | 88 [83–91] | 86 [81–90] | 0.90 [0.86–0.92] | <0.001 |

## DISCUSSION

The primary motivation for this study was to address the persistent diagnostic challenges of glaucoma in low-resource settings, where late presentation, limited access to subspecialty care, and fragmented diagnostic information remain common, by exploring machine learning techniques in glaucoma diagnosis. In many such settings, including much of SSA, glaucoma diagnosis is often based on a combination of routine clinical examination, basic functional testing, and selectively available imaging, rather than on advanced or standalone technologies. This study, therefore, sought to determine whether machine learning models trained on multimodal, clinically realistic data could enhance diagnostic accuracy beyond what is achievable using individual structural or functional parameters.

Our results directly support this premise. While several individual parameters demonstrated moderate to good discriminative ability, no single structural or functional metric matched the performance of the MLP. This finding underscores a central theme of this study, which is that glaucoma is a multifactorial disease, and its reliable detection requires integrative approaches rather than reliance on isolated measurements.

Consistent with prior literature, age alone showed no meaningful diagnostic utility (AUROC = 0.49), despite its epidemiological association with an increased risk of glaucoma [23]. This reinforces the distinction between risk factors and diagnostic markers, particularly in cross-sectional clinical decision-making [24]. Functional indices derived from standard automated perimetry, notably mean deviation (MD, AUROC = 0.75) and pattern standard deviation (PSD, AUROC = 0.74), demonstrated strong diagnostic performance [25], [26]. These findings align with longstanding evidence that visual field loss reflects established glaucomatous damage.

Structural OCT-derived parameters, particularly RNFL and GCC measurements, showed consistently higher AUROC values than optic nerve head metrics. Inferior and superior RNFL sectors, as well as GCC loss indices (FLV and GLV), were among the strongest individual discriminators. These results reflect the known anatomical pattern of glaucomatous damage and corroborate prior reports that macular metrics are sensitive indicators of disease [27], [28]. Nevertheless, even the best-performing individual metrics fell short of the diagnostic performance achieved by the MLP model, highlighting the inherent limitations of univariate decision-making.

Glaucoma rarely presents with uniform changes across all diagnostic domains, particularly in early or atypical cases [29]. By integrating demographic data, routine clinical measures, structural OCT parameters, and functional indices, the models captured complementary and partially independent sources of diagnostic information. This multimodal fusion reflects real-world clinical reasoning, where clinicians synthesize multiple signals to arrive at a diagnosis [30].

Importantly, the superior performance of the MLP suggests that non-linear interactions between features, which may not be fully captured by conventional linear models and simpler analytical approaches, play a critical role in glaucoma classification [31]. This is particularly relevant in African populations, where anatomical variability and disease phenotype may differ from populations from which many diagnostic thresholds were originally derived [32].

Unlike many deep learning studies that rely on raw imaging data, this study intentionally focused on features routinely available in clinical practice. Feature selection was guided not only by statistical relevance but also by clinical plausibility and availability. This approach enhances the translational potential of the model and distinguishes it from highly specialized systems that require expensive hardware or proprietary imaging pipelines. By demonstrating strong performance using selected clinical features rather than raw images, this work provides evidence that effective AI-driven glaucoma detection does not necessarily require deep convolutional architectures or large-scale image datasets. This is particularly important for low-resource settings, where data storage, computational infrastructure, and imaging standardization may be limited.

The performance of our models compares favorably with previously published machine learning studies in glaucoma detection. Reported AUC values in the literature typically range from 0.75 to 0.88 for traditional ML models using structured clinical data, and from 0.85 to 0.95 for deep learning models trained on large imaging datasets [15], [33], [34]. However, many of these studies rely on single-modality inputs and high-end imaging systems that may not be widely accessible [33].

The training and validation curves of the MLP indicate stable convergence with minimal overfitting, supporting the robustness of the model (see Figure 3). The close alignment between training and validation performance suggests that the model learned generalizable patterns rather than dataset-specific noise.

**Figure 3.**
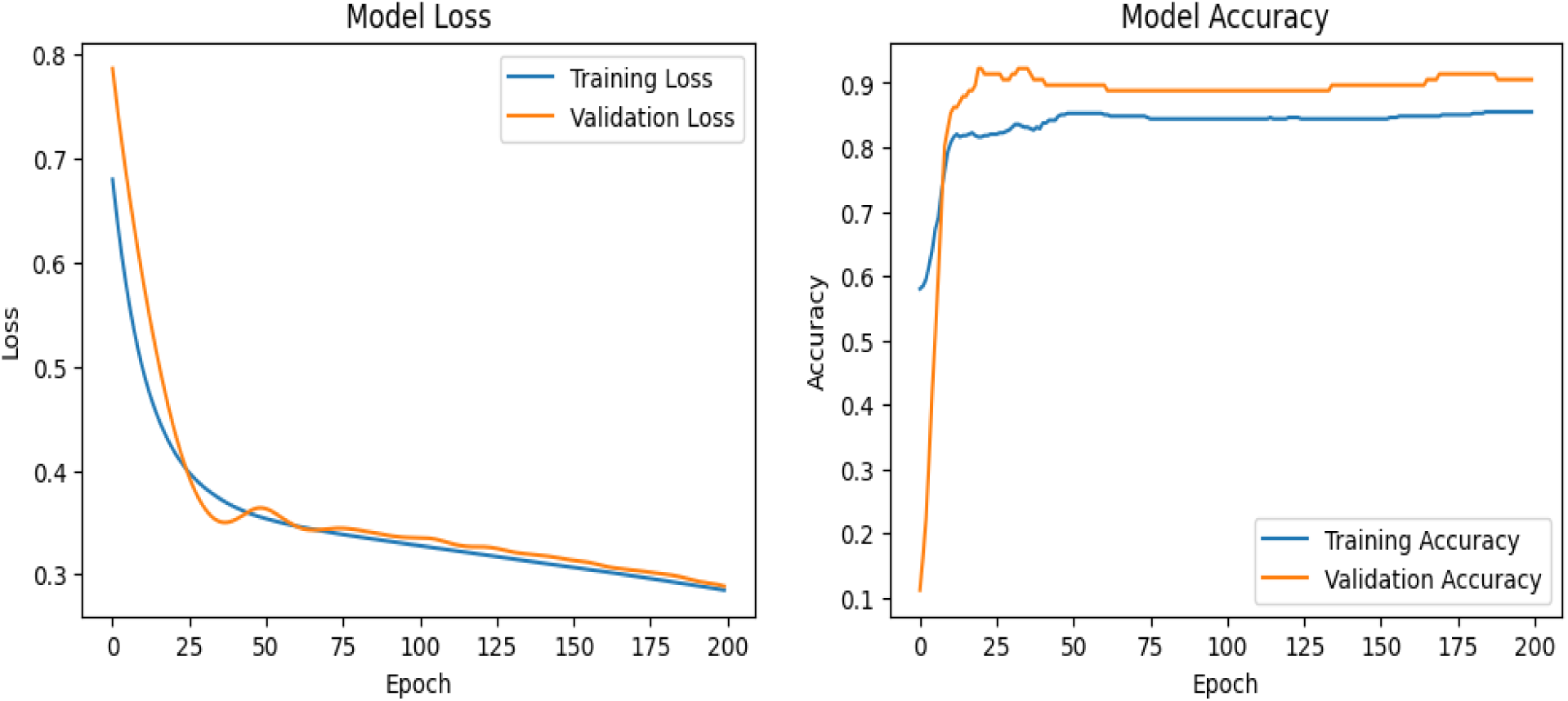
Training and validation curve of the MLP across various epochs. The panel on the left shows progressive reduction in loss, while the right panel demonstrates corresponding improvements in accuracy, with stable convergence and minimal overfitting.

From a clinical perspective, the findings of this study support the potential role of ML-based decision support systems as adjunct tools rather than replacements for clinical judgment. By synthesizing multimodal data into a single probabilistic output, such systems could assist clinicians, particularly non-specialists, in identifying patients who require referral or closer monitoring [35], [36]. At a health systems level, this approach aligns with task-shifting strategies commonly employed in low-resource environments. An ML-based tool embedded within routine clinical workflows could help standardize glaucoma assessment, reduce diagnostic delays, and optimize the use of limited specialist resources.

### Limitations and Future Directions

Despite its strengths, this study has limitations. The sample size and single-population design may limit generalizability; therefore, external validation across diverse clinical settings is needed. Additionally, while the model demonstrated strong performance, future work should incorporate explainability techniques to enhance clinical trust and adoption.

## Conclusion

This study demonstrates that a multimodal machine learning model built with MLP outperforms individual structural and functional metrics in glaucoma diagnosis, particularly within clinically realistic, low-resource settings. By leveraging routinely available clinical data and integrating complementary diagnostic domains, the proposed approach offers a scalable, effective, and context-appropriate pathway toward improved glaucoma detection and care.

### Author Contribution

**Emmanuel Adator:** Conceptualization, Methodology, Project administration, Data Collection, Data curation, Formal analysis, Software, Visualization, Writing – original draft preparation, review & editing. **Andrew Owusu-Ansah:** Conceptualization, Supervision, Methodology, Investigation, Clinical validation, Writing – review & editing, Approval of final draft. **Mercy Oforiwaa Berchie:** Data curation, Analysis, Writing – review & editing. **Julius Markwei:** Investigation, Data analysis, Computational resources, Writing – review & editing. **Joseph Sa-Ambo:** Data acquisition, Investigation, Computational resources. **Anag-Bey Kwame:** Investigation, Computational resources, Data verification. **Abraham Boakye Yiadom:** Computational resources, Writing – review & editing. **Samuel Kyei:** Supervision, Investigation, Clinical resources, Data verification. **Enyam Morny:** Supervision, Investigation, Data verification, Clinical oversight, Writing – review & editing. **Emmanuel Addai**: Investigation, Clinical validation, Project administration, Writing – review & editing.

## Funding details

This study received no external funding.

## Disclosure statement

The authors report there are no competing interests to declare.

## Data availability statement

The authors confirm that the data supporting this study is available upon reasonable request to the corresponding authors and with institutional clearance.

## Clinical trial number

not applicable.

## Notes

### Competing Interest Statement

The authors have declared no competing interest.

### Funding Statement

This study did not receive any funding

### Author Declarations

Ethics committee/IRB of University of Cape Coast, Ghana gave ethical approval for this work.

